# Effectiveness and durability of the mRNA vaccine-induced SARS-CoV-2-specific humoral and cellular immunity in severe asthma patients on biological therapy

**DOI:** 10.1101/2022.02.17.22271122

**Authors:** Michal Podrazil, Pavla Taborska, Dmitry Stakheev, Michal Rataj, Jan Lastovicka, Alena Vlachova, Petr Pohunek, Jirina Bartunkova, Daniel Smrz

**Affiliations:** Department of Immunology, Second Faculty of Medicine, Charles University, and Motol University Hospital, Prague, Czech Republic; Department of Pneumology, Second Faculty of Medicine, Charles University, and Motol University Hospital, Prague, Czech Republic; Department of Pediatrics, Second Faculty of Medicine, Charles University, and Motol University Hospital, Prague, Czech Republic

**Keywords:** severe asthma patients on biological therapy, SARS-CoV-2, COVID-19 vaccination, cellular immunity, humoral immunity

## Abstract

The COVID-19 vaccines effectively elicit humoral and cellular immunity against the severe acute respiratory syndrome coronavirus 2 (SARS-CoV-2) in a healthy population. This immunity decreases several months after the vaccination. However, the efficacy of the vaccine-induced immunity and its durability in patients with severe asthma on biological therapy is unknown. In this study, we evaluated the effectiveness and durability of the mRNA vaccine-induced SARS-CoV-2-specific humoral and cellular immunity in severe asthma patients on biological therapy. The study included 37 patients with severe asthma treated with anti-IgE (omalizumab, n=18), anti-IL5 (mepolizumab, n=14; reslizumab, n=4), or anti-IL5R (benralizumab, n=1) biological therapy. All patients were vaccinated with two doses of BNT162b2 mRNA vaccine (Comirnaty) at a 6-week period between the doses. We found that the COVID-19 vaccination elicited SARS-CoV-2-specific humoral and cellular immunity, which significantly declined 6 months after the second dose of the vaccine. The type of biological treatment did not affect the vaccine-elicited immunity. However, the patients’ age negatively impacted the vaccine-induced humoral response. On the other hand, no such age-related impact was observed on the vaccine-elicited cellular immunity. Our findings showed that biological therapy of patients with severe asthma does not compromise the effectiveness and durability of the COVID-19 vaccine-induced immunity.

## INTRODUCTION

Severe acute respiratory syndrome coronavirus 2 (SARS-CoV-2) is the cause of the COVID-19 pandemic, and the effort in fighting the disease largely relies on prophylactic vaccination (*1*). Currently used COVID-19 vaccines provide a high level of protection against SARS-CoV-2 infection and severe forms of COVID-19 (*2–6*). The COVID-19 vaccines induce a strong humoral and cellular response in a healthy population (*7*). The humoral response leads to the production of anti-SARS-CoV-2 antibodies, which prevent the virus from infecting host cells and spreading in the body (*8, 9*). The cellular response, which largely relies on cytotoxic and memory T cells, prevents the virus from multiplication in the infected cells and protects patients from severe forms of the disease (*10–13*). Despite the high performance of the COVID-19 vaccine in eliciting a robust protective immunity, the durability of this immunity, both humoral and cellular, was found to be much shorter than previously expected (*14–16*). This finding led to a change in the vaccination strategy and application of booster doses to reinvigorate the immunity against the virus (*17*).

Unlike the healthy population, the efficacy of the COVID-19 vaccines in eliciting protective immunity is decreased in patients with many diseases. This has been shown, for instance, in recipients of solid-organ transplants (*18, 19*), cancer patients (*20*), patients on dialysis (*21*), or patients using certain drugs (*22*). In addition to the compromised vaccine’s immunogenicity in these patients, the durability of the elicited immunity is still largely unknown.

Patients with severe asthma on biological treatment were initially expected to be at higher risk of SARS-CoV-2 infection and severity of COVID-19 (*23, 24*). Later studies, however, did not support this expectation (*25–27*). Currently, there is no definite consensus whether these patients and their treatment pose a higher risk towards SARS-CoV-2 infection and severity of the disease than the healthy population, and, therefore, an individualized approach to these patients was recommended (*28*).

A recent study has shown that vaccination of patients with severe asthma on biological therapy with BNT162b2 mRNA vaccine was safe, tolerated, and had minimal impact on the control of their primary disease (*29*). However, the immunogenicity of the vaccine in these patients has not been evaluated.

In the present study, we have evaluated the immunogenicity of the BNT162b2 mRNA (Comirnaty) vaccine and durability of the vaccine-elicited immunity in a cohort of 37 severe asthma patients on biological therapy, which is already a centralized standard of care in the Czech Republic. This cohort included patients on anti-IgE (omalizumab), anti-IL5 (mepolizumab, reslizumab), or anti-IL5R (benralizumab) therapy (*30, 31*). The immunogenicity of the vaccine was evaluated through specific humoral and cellular responses after the first and second doses of the vaccine. The persistence of the elicited immunity was assessed six months after the second dose of the vaccine.

## RESULTS

### COVID-19 vaccination induces high levels of anti-SARS-CoV-2 spike glycoprotein antibodies, which significantly decreases six months after the administration of the second dose of the vaccine

Thirty-seven patients with severe asthma were enrolled in the study. The inclusion criteria were no previous history of COVID-19 or positive tests for SARS-CoV-2 and ongoing management of the patients’ primary disease by biological therapy indicated according to GINA recommendations (*32, 33*). The cohort comprised 18 patients on omalizumab (anti-IgE therapy), 14 patients on mepolizumab (anti-IL5), 4 patients on reslizumab (anti-IL5), and 1 patient on benralizumab (anti-IL5R) therapy. The patients’ median age was 57 years (range 21–73 years), including 22 women and 15 men. The patients’ baseline characteristics before the first COVID-19 vaccine dose are shown in Table 1. All patients were administered two doses of the SARS-CoV-2 spike glycoprotein-based mRNA vaccine BNT162b2 at a 6-week interval between the two doses. We have respected a minimum interval of 48 hours between COVID-19 vaccination and the application of biologics. The samples were obtained within 1 week before the administration of the first and second doses of the vaccine, and then 4 weeks and 6 months after the second dose of the vaccine (Fig. 1A). Eighteen (49%) patients were completely free of any reactions. Nineteen (51%) patients reported commonly described side effects, most of which were classified as very common/common side effects, mostly after the second dose of vaccination. No differences were reported according to the ongoing biologic therapy (data not shown).

**Figure 1.**
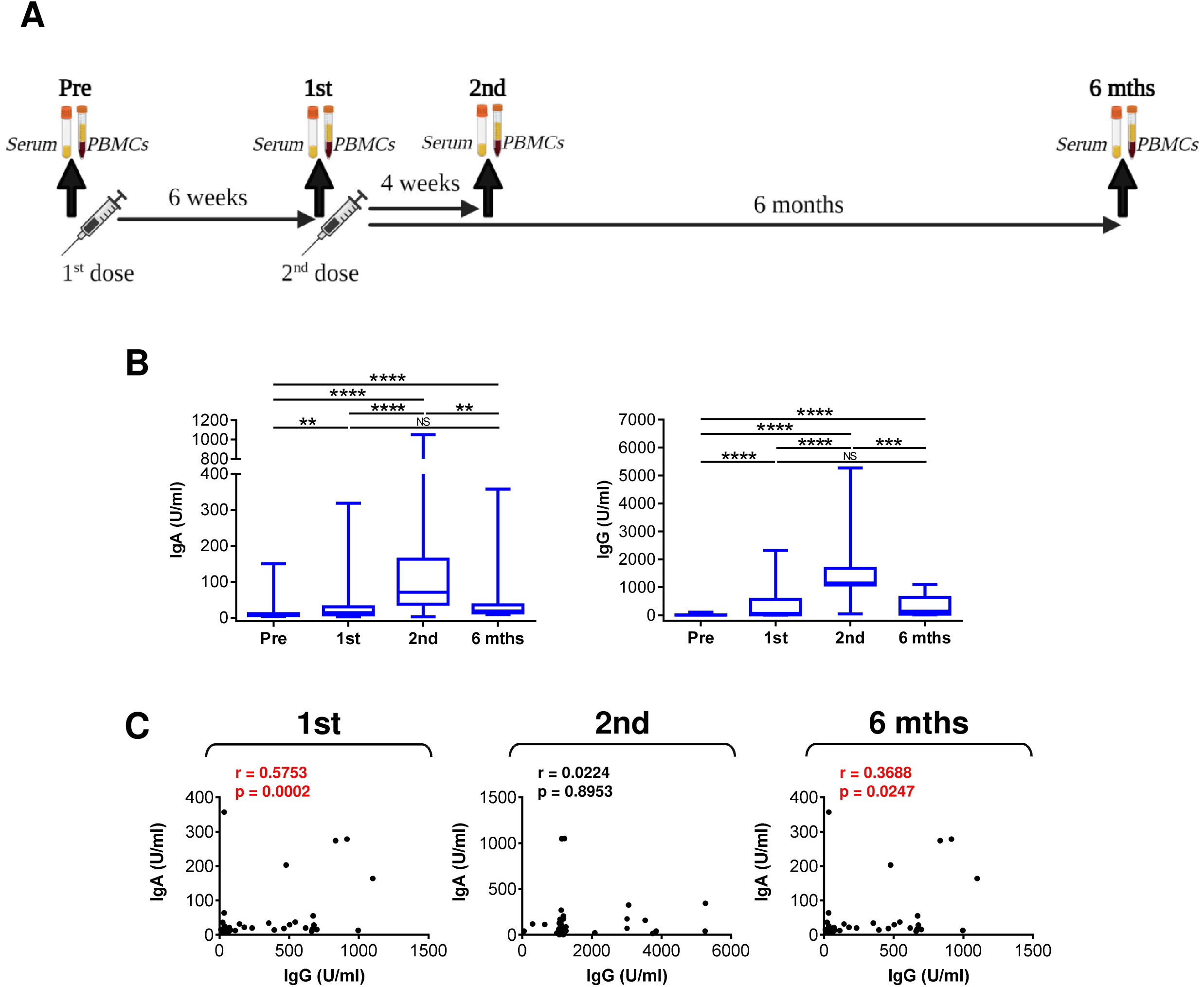
The serum levels of anti-RBD IgA and IgG antibodies during and after the vaccination. (**A**) The sample collection strategy. (**B**) Serum levels (U/ml) of anti-RBD IgA (*left panel*) and IgG (*right panel*) before the first (Pre) and second (1st) vaccine dose, and 4 weeks (2nd) and 6 months (6 mths) after the second vaccine dose. (**C**) The correlations between the serum levels (U/ml) of anti-RBD IgA and IgG antibodies determined in **B**. In **B,** box and whisker plots (2.5–97.5 percentile) and significances of differences among the groups (Pre, 1st, 2nd, and 6 mths) are indicated (^NS^*p>*0.05, \*\**p*<0.01, \*\*\**p*<0.001, \*\*\*\**p*<0.0001; *n* = 37; matched-pair one-way ANOVA with the Dunn’s posttest). In **C,** Spearman’s rank-order correlation coefficient (r) and the significance (*p-*value; *n* = 37) are indicated.

**Table 1:** The cohort characteristics.

| All | Patients<br>(no.) | Patients<br>(%) | Age<br>(median, range) | Race |  |
| --- | --- | --- | --- | --- | --- |
| All | 37 | 100 | 57, 21–73 | Caucasian |  |
| Women | 22 | 59.5 | 55, 21–71 | Caucasian |  |
| Men | 15 | 40.5 | 57, 21–73 | Caucasian |  |
|  | All | Omalizumab<br>(anti-IgE) | Mepolizumab<br>(anti-IL5) | Reslizumab<br>(anti-IL5) | Benralizumab<br>(anti-IL5R) |
| Patients<br>(no.) | 37 | 18 | 14 | 4 | 1 |
| Treatment<br>(months)<br>(median, range) | 34, 1–146 | 73, 17–146 | 20, 1–44 | 33, 30–34 | 9 |
| BMI<br>(median, range) | 26, 17–41 | 25, 17–35 | 27, 20–41 | 26, 21–30 | 27 |
| Blood<br>eosinophils<br>(cells/μl)<br>(median, range) | 110, 0–990 | 165, 30–990 | 45, 20–290 | 135, 50–210 | 0 |
| Total IgE<br>(IU/ml)<br>(median, range) | 231, 17–3067 | 254, 17–799 | 132, 20–3067 | 173, 39–293 | 606 |
| ECP<br>(ng/ml)<br>(median, range) | 12, 3–150 | 12, 3–150 | 12, 4–68 | 18, 8–28 | 10 |
| FeNO<br>(ppb)<br>(median, range) | 43, 5–232 | 36, 5–232 | 64, 6–158 | 86, 35–114 | 43 |

We first evaluated whether the enrolled patients had been infected with SARS-CoV-2 before or during the study to eliminate interference with the vaccine performance. The marker of a previous SARS-CoV-2 infection is the presence of anti-SARS-CoV-2 nucleocapsid protein (NCP) IgG antibodies in serum (*34*). We found that before the vaccination and 6 months after the vaccination 33 patients were negative for anti-NCP IgG antibodies (Fig. S1A). Among these negative 33 patients, we found only two patients with the pre-vaccination-elevated anti-SARS-CoV-2 spike glycoprotein receptor-binding domain (RBD) IgA antibodies and one patient with the pre-vaccination-elevated anti-RBD IgG antibodies (Fig. S1B). The remaining 4 patients of the cohort had low or borderline levels of anti-NCP IgG antibodies, indicating a distant SARS-CoV-2 infection in the past (Fig. S1A). Regardless, all four patients were negative for anti-RBD IgA and IgG antibodies (Fig. S1C). These data showed that the patients’ possible SARS-CoV-2 infection had not largely compromised the evaluation of the COVID-19 vaccine performance during the study implementation.

To determine the COVID-19 vaccine performance in eliciting a humoral response in the tested patients, we analyzed serum levels of anti-RBD IgA and IgG antibodies during and after the vaccination. As shown in Fig. 1B, the serum levels of anti-RBD IgA and IgG antibodies significantly increased after the first dose of the vaccine. These serum levels further and significantly increased after the second dose of the vaccine (Fig. 1B). However, 6 months after the second dose of the vaccine, these levels declined back to levels observed after the first dose (Fig. 1B). As shown in Fig. 1C, the serum levels of anti-RBD IgA and IgG antibodies were found to correlate after the first and 6 months after the second dose of the vaccine but not 4 weeks after the second dose. This finding indicated that the second dose of the vaccine more promoted the IgG antibodies than IgA antibodies (Fig. 1C). Collectively, the data showed that the immunogenicity of BNT162b2 vaccine is sufficient to induce a strong humoral immunity in severe asthma patients on biological therapy but that this immunity then significantly declines 6 months after the second dose of the vaccine.

### COVID-19 vaccine induces SARS-CoV-2 spike glycoprotein-specific CD4^+^ and CD8^+^ T cell immunity, which significantly decreases six months after the administration of the second dose of the vaccine

The cellular immunity against SARS-CoV-2 is important for efficient protection against the virus (*35*). Unlike humoral immunity, where antibodies immediately neutralize the virus, cellular immunity needs to mobilize against the virus after the antigen challenge (*36–38*). This mobilization relies on both CD4^+^ and CD8^+^ T cells to recognize the target antigen and their effective proliferation after the antigen recognition. To exclude that the patients’ biological therapy compromised both these abilities of T cells, we tested the cellular immunogenicity of the vaccine using peptides derived from SARS-CoV-2 spike glycoprotein (*39, 40*). These peptides were used to *in vitro* enrich the patients’ PBMCs with the peptide-reactive TNFα-, IFNγ- or TNFα/IFNγ-producing CD4^+^ or CD8^+^ T cell populations (Fig. 2A).

**Figure 2.**
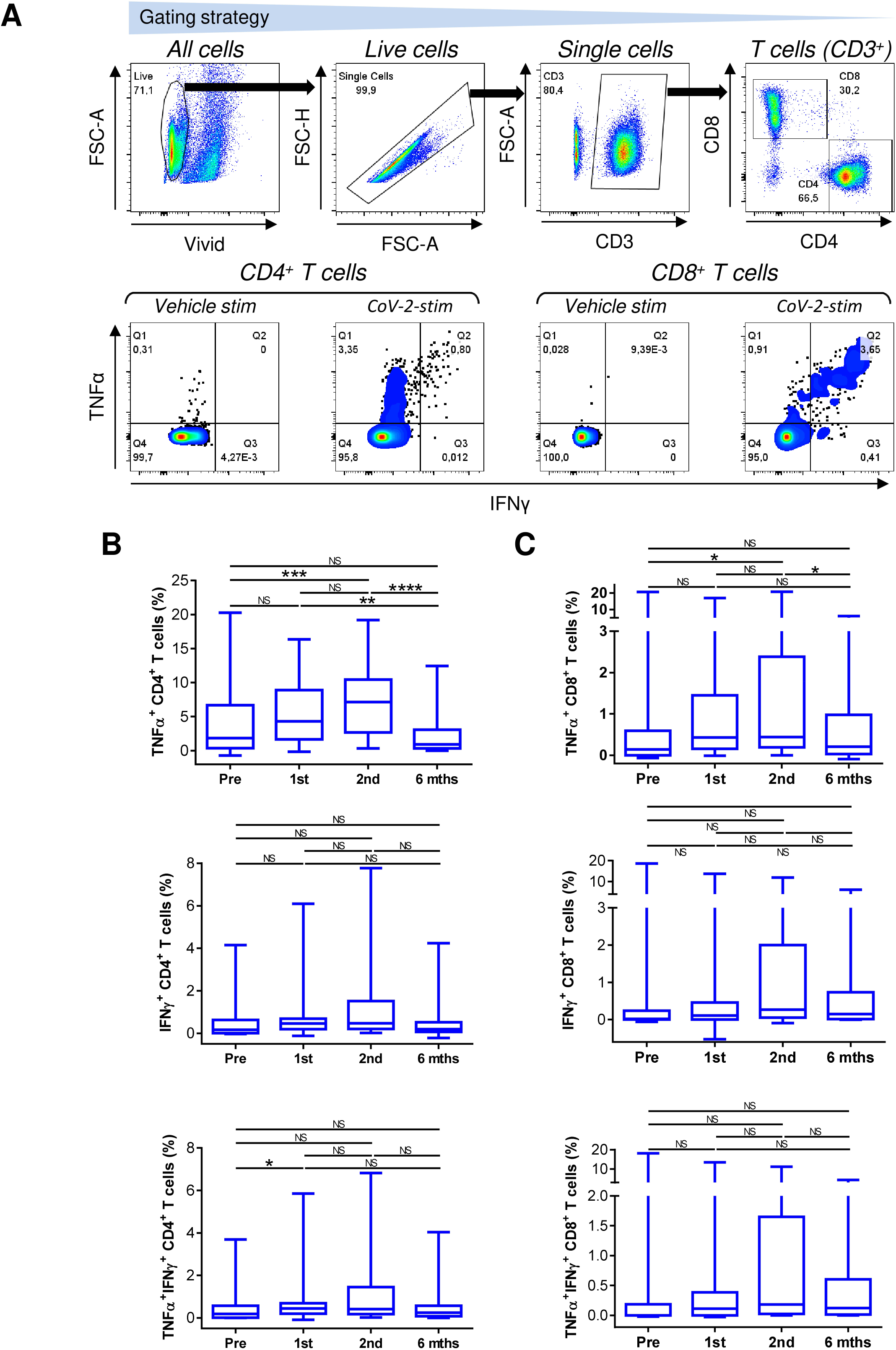
Reactivity of the enriched PBMCs to SARS-CoV-2 spike glycoprotein-peptides. (**A**) The gating strategy of flow cytometry data (*top panels*). The frequency of TNFα-, IFNγ-, or TNFα/IFNγ-producing CD4^+^ and CD8^+^ T cells determined by intracellular cytokine staining in the enriched PBMCs stimulated with vehicle (Vehicle-stim) or SARS-CoV-2 spike glycoprotein-derived peptides (CoV-2-stim) (*bottom panel*s). The frequency of peptide-reactive cytokine-producing T cell populations was calculated as the difference between the frequency of the cytokine-producing T cells in the vehicle- and the peptide-stimulated sample. (**B**–**C**) The frequency of peptide-reactive TNFα-, IFNγ-, or TNFα/IFNγ-producing CD4^+^ (**B**) and CD8^+^ (**C**) T cells before the first (Pre) and second (1st) vaccine dose, and 4 weeks (2nd) and 6 months (6 mths) after the second vaccine dose. In **B**–**C,** box and whisker plots (2.5–97.5 percentile) and significances of differences among the groups (Pre, 1st, 2nd, and 6 mths) are indicated (^NS^*p>*0.05, \**p*<0.05, \*\**p*<0.01, \*\*\**p*<0.001, \*\*\*\**p*<0.0001; *n* = 37; matched-pair one-way ANOVA with the Dunn’s posttest).

We found that the COVID-19 vaccination enhanced the mobilization of SARS-CoV-2-reactive TNFα-producing CD4^+^ and CD8^+^ T cells and this enhancement was significant after the second dose of the vaccine (Fig. 2B and 2C, top panels). However, this enhanced mobilization significantly declined to pre-vaccination levels 6 months later (Fig. 2B).

The mobilization of SARS-CoV-2-reactive IFNγ- or TNFα/IFNγ-producing CD4^+^ or CD8^+^ T cell populations showed a similar tendency as in the TNFα-producing T cells, but the data did not reach statistical significance within the size of the tested cohort (Fig. 2B and 2C, middle and bottom panels). However, the data showed that the cellular mobilization of the reactive cytokine-producing CD4^+^ and CD8^+^ T cells significantly correlated (Fig. 3), indicating polyfunctional responses of the patients’ SARS-CoV-2-reactive T cells after the antigen challenge. These data showed that the immunogenicity of BNT162b2 vaccine is sufficient to also significantly promote cellular immunity in severe asthma patients on biological therapy but that this immunity, similar to the humoral immunity, significantly declines 6 months after the second dose of the vaccine.

**Figure 3.**
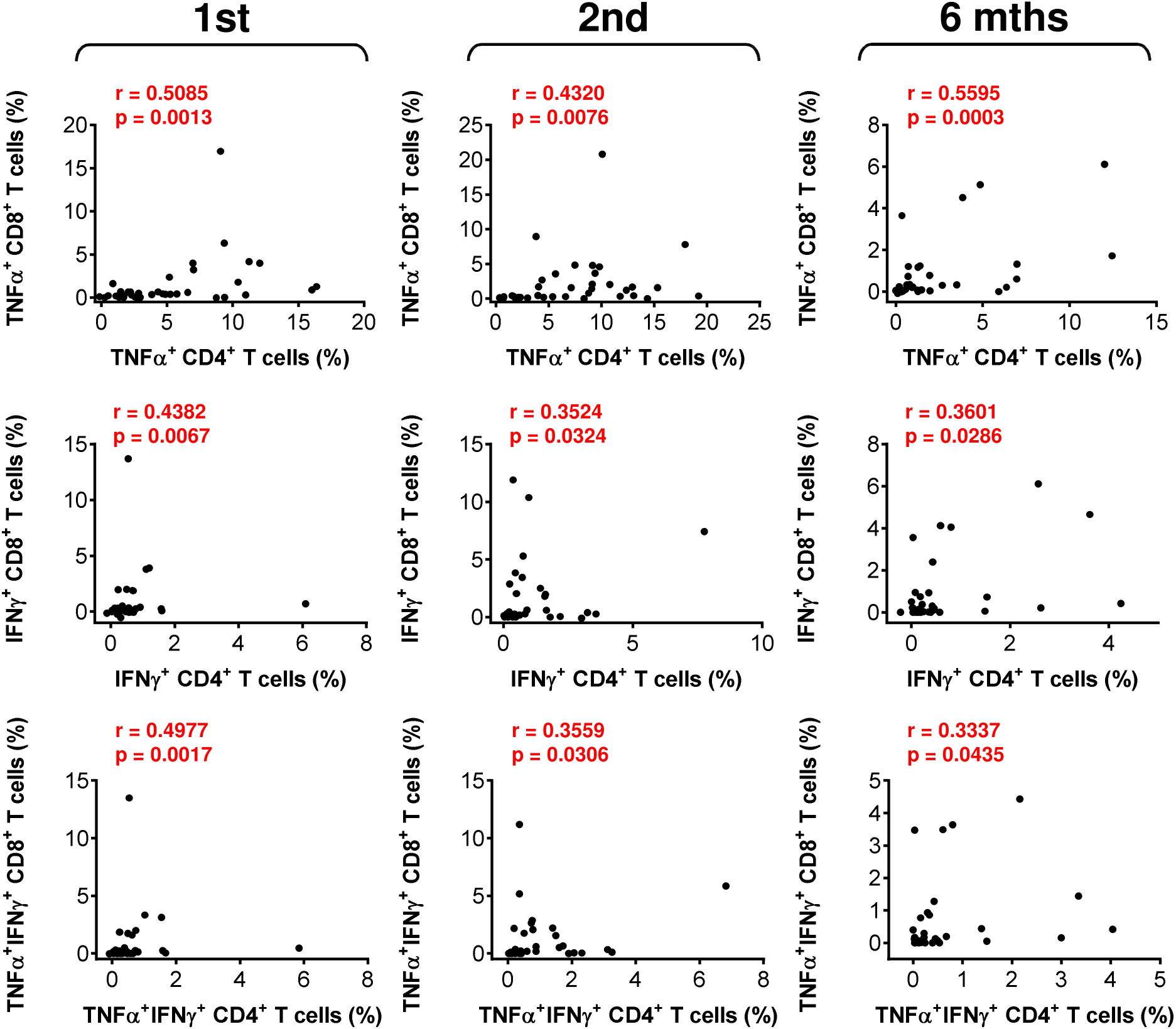
The association between SARS-CoV-2 spike glycoprotein peptide reactive CD4^+^ and CD8^+^ T cell reactivity during and after the vaccination. The correlations between the frequency of peptide-reactive TNFα-, IFNγ-, or TNFα/IFNγ-producing CD4^+^ and CD8^+^ T cells before the second (1st) vaccine dose, and 4 weeks (2nd) and 6 months (6 mths) after the second vaccine dose. The Spearman’s rank-order correlation coefficient (r) and the significance (*p*-value; *n* = 37) are indicated.

### COVID-19 vaccine-induced humoral immunity correlates with SARS-CoV-2-specific CD8^+^ T cell immunity

The data showed that the COVID-19 vaccine induced both humoral and cellular immunity in severe asthma patients on biological therapy. Next, we analyzed whether there was a correlation between the extent of the vaccine-elicited humoral and cellular immunity. We found that serum levels of anti-RBD IgA and IgG antibodies minimally correlated with SARS-CoV-2-reactive CD4^+^ T cells (Fig. S2). On the other hand, these antibody serum levels significantly correlated with SARS-CoV-2-reactive CD8^+^ T cells (Fig. 4). These findings showed that the COVID-19 induced SARS-CoV-2-specific humoral immunity is proportionally translated into the CD8^+^ T cell counterpart.

**Figure 4.**
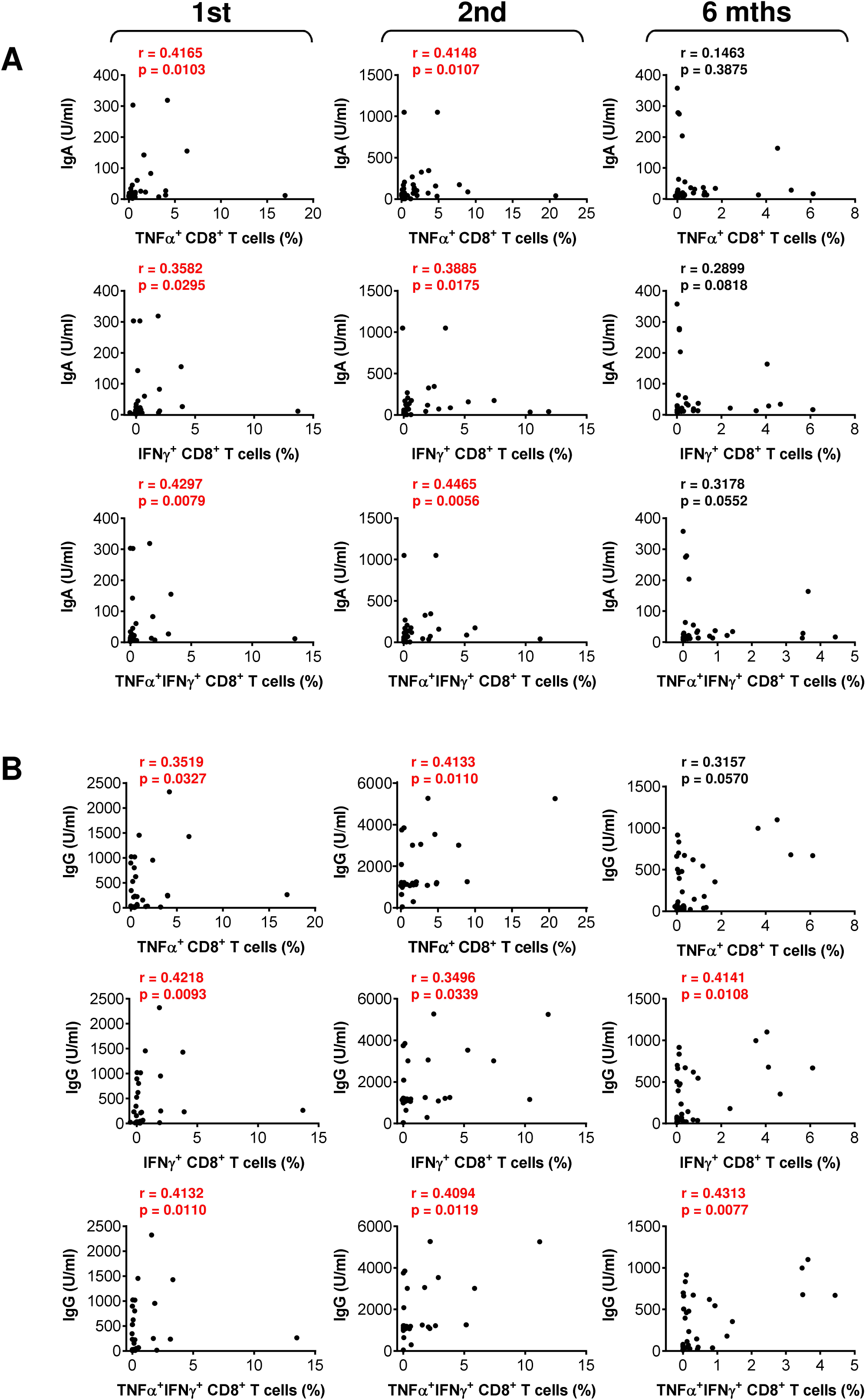
The association between SARS-CoV-2 spike glycoprotein peptide reactive CD8^+^ T cells and serum levels of anti-RBD antibodies during and after the vaccination. (**A**–**B**) The correlations between the frequency of peptide-reactive TNFα-, IFNγ-, or TNFα/IFNγ-producing CD8^+^ T cells and serum levels (U/ml) of anti-RBD IgA (**A**) and IgG (**B**) antibodies before the second (1st) vaccine dose, and 4 weeks (2nd) and 6 months (6 mths) after the second vaccine dose. The Spearman’s rank-order correlation coefficient (r) and the significance (*p*-value; *n* = 37) are indicated.

### Immunogenicity of the COVID-19 vaccine is comparable in severe asthma patients on different biologicals

The COVID-19 vaccine effectively induced SARS-CoV-2-specific immunity in the studied cohort of 37 patients on biological therapy. We next analyzed whether the type of biological therapy impacted the vaccine performance. We stratified the patients into 3 groups based on the therapy type. The first group included 18 patients on anti-IgE therapy (omalizumab), the second group 14 patients on anti-IL5 therapy (mepolizumab), and the third group 4 patients on anti-IL5 (reslizumab). One patient on a different drug, anti-IL5R therapy (benralizumab), was excluded from these comparisons. In the stratified 3 groups, there were no significant differences in the age of the patients, their total IgE or eosinophil cationic protein (ECP) serum levels (Fig. S3). The only difference in the tested parameters among the groups was the eosinophil counts in peripheral blood between the group treated with anti-IgE therapy (omalizumab) and the group treated with anti-IL5 therapy (mepolizumab) (Fig. S3).

We found that the type of biological therapy had no significant impact on the COVID-19 vaccine-elicited humoral immunity (Fig. 5A). No impact was also found on the vaccine-elicited cellular immunity (Fig. 5B). These findings revealed that the type of the biological therapy showed no adverse effect on the vaccine’s performance in the treated patients with severe asthma.

**Figure 5.**
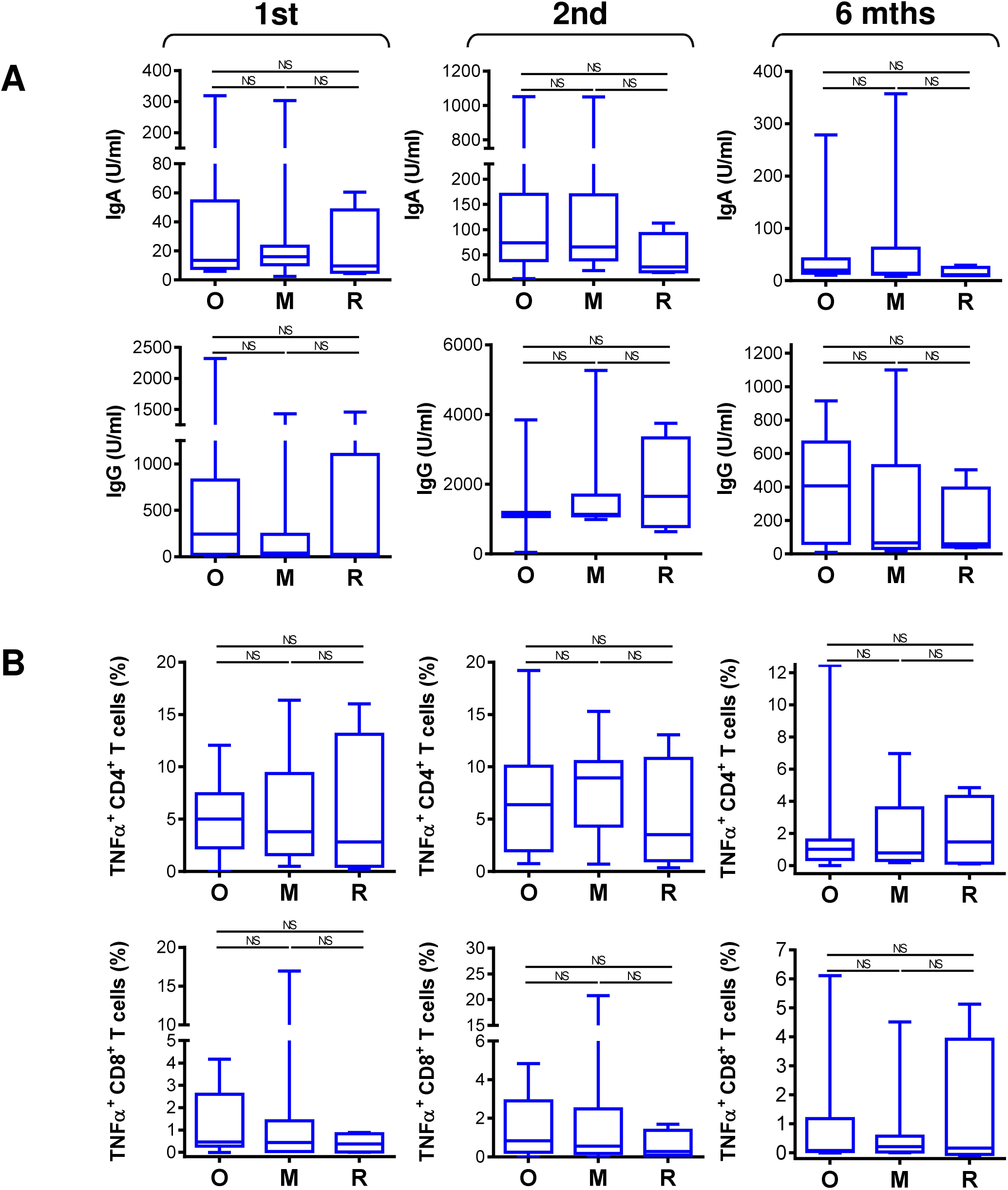
The impact of biological therapy on SARS-CoV-2 spike glycoprotein-specific humoral and cellular immunity during and after the vaccination. (**A**) The patients were stratified into 3 groups based on the type of biological therapy. The groups were 18 patients on anti-IgE (O; omalizumab), 14 patients on anti-IL5 (M, mepolizumab), and 4 patients on anti-IL5 (R, reslizumab) therapy. Serum levels (U/ml) of anti-RBD IgA (*top panels*) and IgG (*bottom panels*) before the second (1st) vaccine dose, and 4 weeks (2nd) and 6 months (6 mths) after the second vaccine dose. (**B**) The frequency of peptide-reactive TNFα-producing CD4^+^ (*top panels*) and CD8^+^ (*bottom panels*) T cells in the stratified groups in **A** before the second (1st) vaccine dose, and 4 weeks (2nd) and 6 months (6 mths) after the second vaccine dose. In (**A**–**B**), box and whisker plots (2.5–97.5 percentile) and significances of differences among the groups (O, M, R) are indicated (^NS^*p>*0.05; *n* = 18 (O), 14 (M), or 4 (R); matched-pair one-way ANOVA with the Dunn’s posttest).

### COVID-19 vaccine-induced humoral but not cellular immunity is negatively affected by the age of the severe asthma patients on biological therapy

So far, we found no clinical parameter that would negatively affect the COVID-19 vaccine performance in the studied cohort of severe asthma patients. We next analyzed whether any of the other clinical parameters affected the vaccine-elicited humoral or cellular immunity. We found that total IgE or ECP serum levels did not correlate with the vaccine-elicited humoral (Fig. S4) or cellular immunity (Fig. S5 and S6). Also, the peripheral blood eosinophil counts did not correlate with the vaccine-elicited CD8^+^ T cell immunity (Fig. S7). However, eosinophil counts correlated with serum levels of the vaccine-elicited anti-RBD IgG antibodies after the first and 6 months after the second dose of the vaccine (Fig. S8A). After the first vaccine dose, the eosinophil counts also correlated with the vaccine-elicited CD4^+^ T cell immunity (Fig. S8B). Regardless of these findings, we revealed that eosinophil counts correlated with the age of the patients (Fig. S8C). This finding suggested that their age could be the clinical parameter that impacted the vaccine performance. Indeed, the analyses revealed that the age of the patients negatively affected serum levels of the vaccine-elicited anti-RBD IgA and IgG antibodies after the first dose of the vaccine (Fig. 6A). The age of the patients also negatively impacted serum levels of the vaccine-elicited anti-RBD IgG antibodies 6 months after the second dose (Fig. 6A). On the other hand, the age of the patients did not negatively impact the vaccine-elicited cellular immunity (Fig. 6B and Fig. S9). These data showed that the vaccine-elicited humoral immunity was more sensitive to the age of the patients than its cellular counterpart.

**Figure 6.**
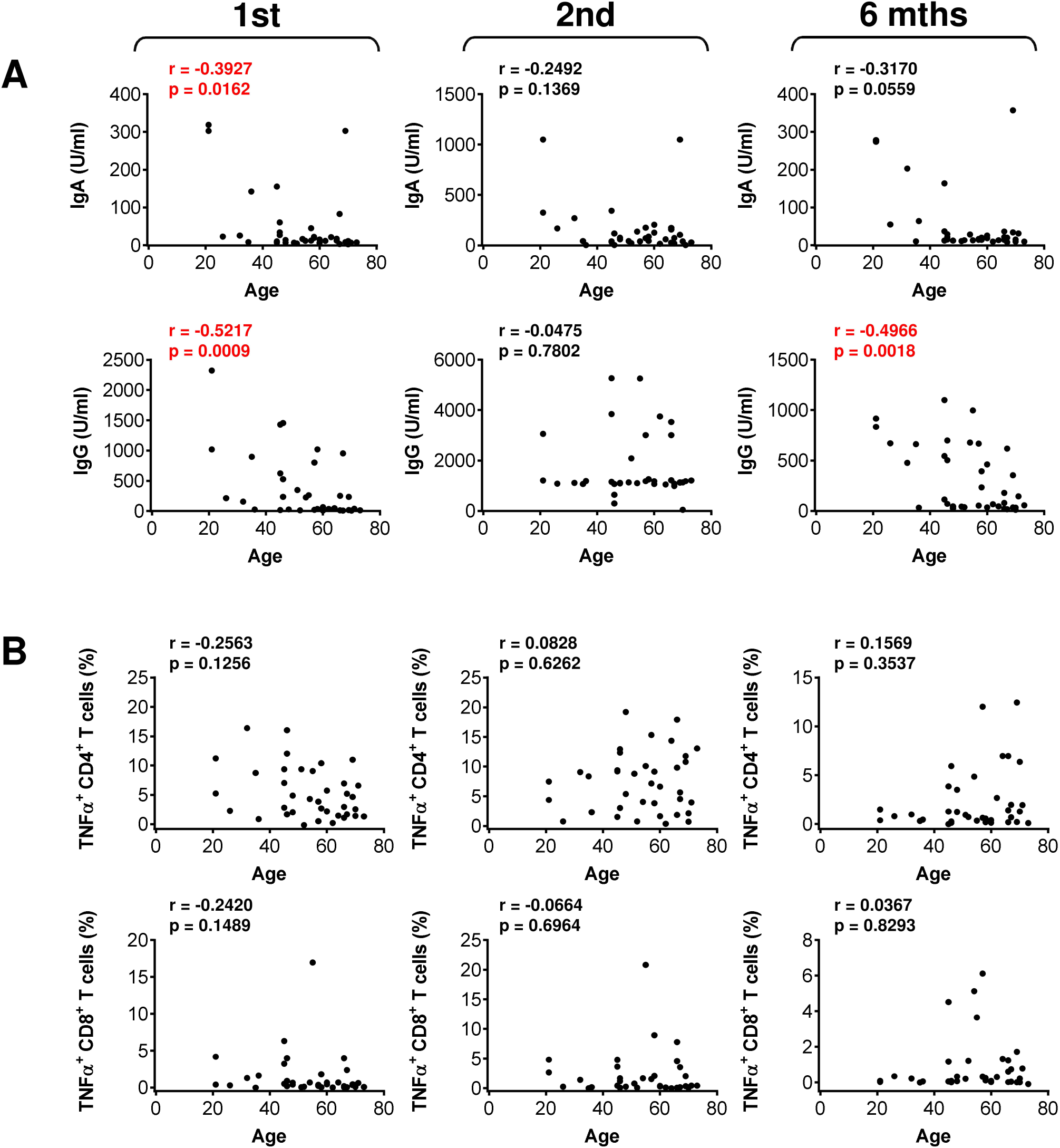
The impact of the patients’ age on serum levels of anti-RBD antibodies and SARS-CoV-2 spike glycoprotein peptide-reactive CD4^+^ and CD8^+^ T cells during and after the vaccination. (**A**) The correlations between the patients’ age and serum levels (U/ml) of anti-RBD IgA (*top panels*) and IgG (*bottom panels*) antibodies before the second (1st) vaccine dose, and 4 weeks (2nd) and 6 months (6 mths) after the second vaccine dose. (**B**) The correlations between the patients’ age and the frequency of peptide-reactive TNFα-producing CD4^+^ (*top panels*) and CD8^+^ (*bottom panels*) T cells before the second (1st) vaccine dose, and 4 weeks (2nd) and 6 months (6 mths) after the second vaccine dose. In (**A**–**B**), Spearman’s rank-order correlation coefficient (r) and the significance (*p*-value; *n* = 37) are indicated.

## DISCUSION

This study showed that the COVID-19 vaccine could elicit both humoral and cellular immunity against the virus in patients with severe asthma on biological therapy. The type of biological therapy had no impact on both arms of the vaccine-elicited immunity. However, similar to the healthy population, the vaccine-elicited immunity significantly decreased 6 months after the vaccination.

Biological therapy is increasingly used in clinical practice in severe asthma patients (*41*). Apart from treating the disease, it impacts the immune system. Patients with severe asthma are often treated with biologicals, which are strong immunomodulators. Omalizumab binds to free IgE antibodies, which lowers free IgE levels and causes FcεRI receptors on basophils and mast cells to be downregulated (*30*). IgE antibodies are also important for cross-talk between innate and adaptive immunity and play a role in susceptibility to respiratory infections (*42*) and antiviral responses (*43*). As such, anti-IgE monoclonal antibodies were suggested as a potential treatment in COVID-19 to enhance the antiviral responses to the virus (*44*). Mepolizumab and reslizumab (anti-IL5 therapy) bind to IL-5 and therefore stop IL-5 from binding to its receptor on the surface of eosinophils. Inhibiting IL-5 binding to eosinophils then reduces blood, tissue, and sputum eosinophil levels. Benralizumab specifically binds to IL-5Rα, thereby preventing the interaction between IL-5 and its receptor. Simultaneously, benralizumab, through its Fc constant region, binds to the FcγIIIRa receptor expressed by natural killer cells, thus inducing eosinophil apoptosis operated by the release of proapoptotic proteins such as granzymes and perforins (*31*). However, even though anti-IL5 therapy decreases the counts of eosinophils, this decrease does not prevent activation of the remaining eosinophils and subsequent humoral and cellular response to viral challenge (*45*). Similar results were observed with anti-IL5R therapy, where the risk of respiratory infections due to decreased eosinophil counts was not largely increased (*46, 47*). Regardless of the different and potent mechanisms through which these anti-type 2 inflammation biological drugs modulate the immune system, these drugs were shown to pose no increased risk of SARS-CoV-2 infection nor worsen the clinical course and outcome of COVID-19 (*48*). The results of our study showed that these drugs also did not prevent the patients’ immune system from eliciting SARS-CoV-2 antigen-specific humoral and cellular immunity after the vaccination. In addition, the durability of this immunity was comparable to the durability of the vaccine-elicited immunity in a healthy population (*14–16*). Therefore, biological drugs used to treat patients of this study are different from immunomodulatory drugs, which were shown to compromise the vaccine’s immunogenicity (*49–51*). On the other hand, these drugs are comparable to immunomodulatory drugs, which were found not to compromise the vaccine’s immunogenicity (*51, 52*).

The COVID-19 vaccine has been shown to elicit SARS-CoV-2-specific immunity in multiple studies, and this capability relates to both humoral and cellular immunity (*2–6*). The dynamics of the humoral response and its durability in patients of our study were well comparable to the data obtained in healthy volunteers (*53*), indicating that the ongoing biological therapy of the patients had no apparent impact on the dynamics and durability of the vaccine-elicited immunity. Our data also showed that, comparably to other studies (*53*), the vaccine promoted more CD4^+^ T than CD8^+^ T cell SARS-CoV-2-specific immunity (*40, 54*). And also, comparably to other reports, the extent of the SARS-CoV-2-specific CD4^+^ and CD8^+^ T cell immunity correlated (*7*). In addition, the extent of humoral immunity correlated with the T cell immunity, namely with the CD8^+^ T cell one. These data, therefore, showed that the vaccine proportionally induces different arms of the immune system in severe asthma patients on biological therapy. However, whether biological therapy impacts this proportionality needs to be further investigated.

The immunogenicity of mRNA vaccines decreases with the age of the vaccinated subjects (*55, 56*). This decrease relates to both the vaccine-elicited humoral and cellular immunity (*55*). We also found a strong correlation between the vaccine-elicited humoral immunity and the age of the vaccinated patients after the first dose of the vaccine and then six months after the second dose. These findings indicate that the age of the severe asthma patients on biological therapy is a negative predictor of the onset and durability of the vaccine-elicited humoral immunity. Surprisingly, the age of the patients not too much affected the SARS-CoV-2-specific cellular immunity. However, it should be noted that the study did not include patients over the age of 80 years, in whom the decline was found to be most pronounced (*55*). Regardless, the age of the severe asthma patients played a significant role in eliciting the humoral immunity and, similarly to the healthy population, needs to be considered as an indicator for the third booster vaccine dose, which was found to significantly restore sufficient SARS-CoV-2-specific immunity (*57*).

This study showed that patients with severe asthma on biological therapy could elicit both arms of SARS-CoV-2-specific immunity after two-dose vaccination with the BNT162b2 mRNA vaccine. However, comparable to the healthy population, this immunity is not presumably sufficient 6 months after the vaccination, and a booster dose should be considered.

## MATERIALS AND METHODS

### Patients and COVID-19 vaccination

The study included 37 patients with severe asthma who were on biological therapy and previously planned a voluntary vaccination against COVID-19 with the BNT162b2 mRNA SARS-CoV-2 vaccine (Pfizer-BioNTech). The blood samples were obtained between April 2021 and November 2021. The planned COVID-19 vaccination included 2 doses of the BNT162b2 SARS-CoV-2 vaccine (Pfizer-BioNTech) administered at a 6-week interval between each dose. According to the ongoing treatment with monoclonal antibodies for severe asthma, a 48-h interval between COVID-19 vaccination and the biologics was used. The patients’ samples for the analyses included peripheral blood serum and unclotted peripheral blood and were collected during the planned health check-ups. Complete blood count with differential was performed. The serum and peripheral blood mononuclear cells (PBMCs) were isolated and cryopreserved as previously described (*40, 58*). Adverse reactions to mRNA vaccination were monitored. Each patient provided signed written informed consent for the use of their blood-derived products for future research. The study was approved according to the ethical standards of the institutional research committee – the Ethics Committee of the Motol University Hospital in Prague (EK-346/21), and performed in compliance with the 1964 Helsinki declaration and its later amendments or comparable ethical standards.

### Determination of the serum levels of anti-SARS-CoV-2 antibodies

The patients’ sera were analyzed for the presence of anti-SARS-CoV-2 antibodies. Anti-SARS-CoV-2-spike glycoprotein receptor-binding domain (RBD) IgA and IgG antibodies were determined using IVD EIA COVID-19 RBD IgA or IgG (TestLine Clinical Diagnostics, Brno, Czech Republic). Anti-SARS-CoV-2 nucleocapsid protein (NCP) IgG antibodies were determined using CLIA COVID-19 NP IgG (TestLine Clinical Diagnostics). The analyses were performed according to the manufacturer’s instructions. The levels of the specific antibodies were expressed according to the manufacturer’s instructions in (U/ml). The samples with (U/ml) values below 18 were considered negative, and levels above 18 and more were judged as positive. Samples with antibody levels above the dynamic range of the method were reanalyzed using an adequately diluted serum.

### Enrichment of patients’ PBMCs with SARS-CoV-2-reactive T Cells

The cryopreserved PBMCs were processed with minor changes as previously described (*40*). Briefly, the cells were reconstituted (2 × 10^6^ cells/ml) overnight in a human plasma serum-containing medium and then stimulated with overlapping peptides (0.5 μg/ml) of SARS-CoV-2 spike glycoprotein [PepMix™ SARS-CoV-2 Spike Glycoprotein, cat.# PM-WCPV-S-1, JPT Peptide Technologies, Berlin, Germany]. The stimulated cells were then cultured (37 °C, 5% CO_2_) for 8 days in the presence of IL-2 (Peprotech, Cranbury, NJ) to enrich the cells with peptide-reactive T cells.

### SARS-CoV-2 spike glycoprotein peptide reactivity of the enriched PBMCs

The 8-day-enriched PBMCs were restimulated with SARS-CoV-2 spike glycoprotein-derived peptides (0.5 μg/ml) and cultured for 5 h in the presence of brefeldin A (BioLegend, San Diego, CA) supplemented 1 h after the stimulation. The samples stimulated with vehicle alone (the peptide solvent) were used as a control. After the stimulation, the cells were stained with fixable live/dead stain, fixed, permeabilized, and stained with fluorescent-tagged CD3-, CD4-, CD8-, IFN-γ-, and TNF-α-specific antibodies as described (*40*). The stained cells were analyzed by flow cytometry (FACSAria II, Becton Dickinson, Heidelberg, Germany). FlowJo software (Tree Star, Ashland, OR) was used to analyze the acquired flow cytometry data. The percentage of the peptide-reactive cytokine-producing T cell populations was calculated as the difference between the percentage of the cytokine-producing T cells in the vehicle- and the peptide-stimulated sample.

### Statistical Analysis

The values were calculated from the indicated sample size (*n*) using GraphPad Prism 6 (GraphPad Software, La Jolla, CA). Wilcoxon matched-pair signed-rank test was used to calculate the statistical significance (\**p*<0.05, \*\**p*<0.01, \*\*\**p*<0.001, \*\*\*\**p*<0.0001) between two variables. The matched-pair one-way ANOVA with Dunn’s posttest was used to determine the statistical significance (^NS^*p>*0.05, \**p*<0.05, \*\**p*<0.01, \*\*\**p*<0.001, \*\*\*\**p*<0.0001) between three or more variables. The Spearman’s rank-order correlation coefficient (*r*) and the correlation’s statistical significance (*p*) were used to determine the associations between two variables. The *p*-value below 0.05 was considered significant. Biorender.com was used to produce graphical images (accessed in January 2022).

## Data Availability

All data produced in the present study are available upon reasonable request to the authors.

## ACKNOWLEDGMENTS

Research in the authors’ laboratories was supported by the institutional IPE2 funding of the Charles University, Second Faculty of Medicine in Prague.

## Author contributions

M.P., P.T., D.S., and Da.S. designed the experiments. P.T., D.S., M.R, J.L., and Da.S. conducted the experiments and/or analyzed the data. M.P., A.V., P.P., and J.B. supervised the sample collection and clinical aspects of the study; M.P. and Da.S. wrote the manuscript; P.T., D.S., M.R, J.L., A.V., P.P., and J.B. contributed to the writing of the manuscript; M.P. and Da.S. supervised the research.

## Conflict of interest

J.B. is a part-time employee and a minority shareholder of Sotio, a.s. P.P. is a member of the advisory board of the GSK and received a consultancy fee from AstraZeneca, Novartis, Chiesi and speakeŕs fee from GSK and Novartis. M.P., P.T., D.S., M.R, J.L., A.V., and Da.S. declare no conflicts of interest.

## Ethical statement

All experimental protocols were approved by the ethical standards of the institutional national research committee – the Ethics Committee of the Motol University Hospital in Prague (protocol no. EK-346/21). All experiments were performed in accordance with the 1964 Helsinki declaration and its later amendments or comparable ethical standards. All patients included in the study signed informed consent for the use of their blood-derived products for future research.

## SUPPLEMENTARY MATERIAL

### SUPPLEMENTARY FIGURE LEGENDS

**Figure S1.**
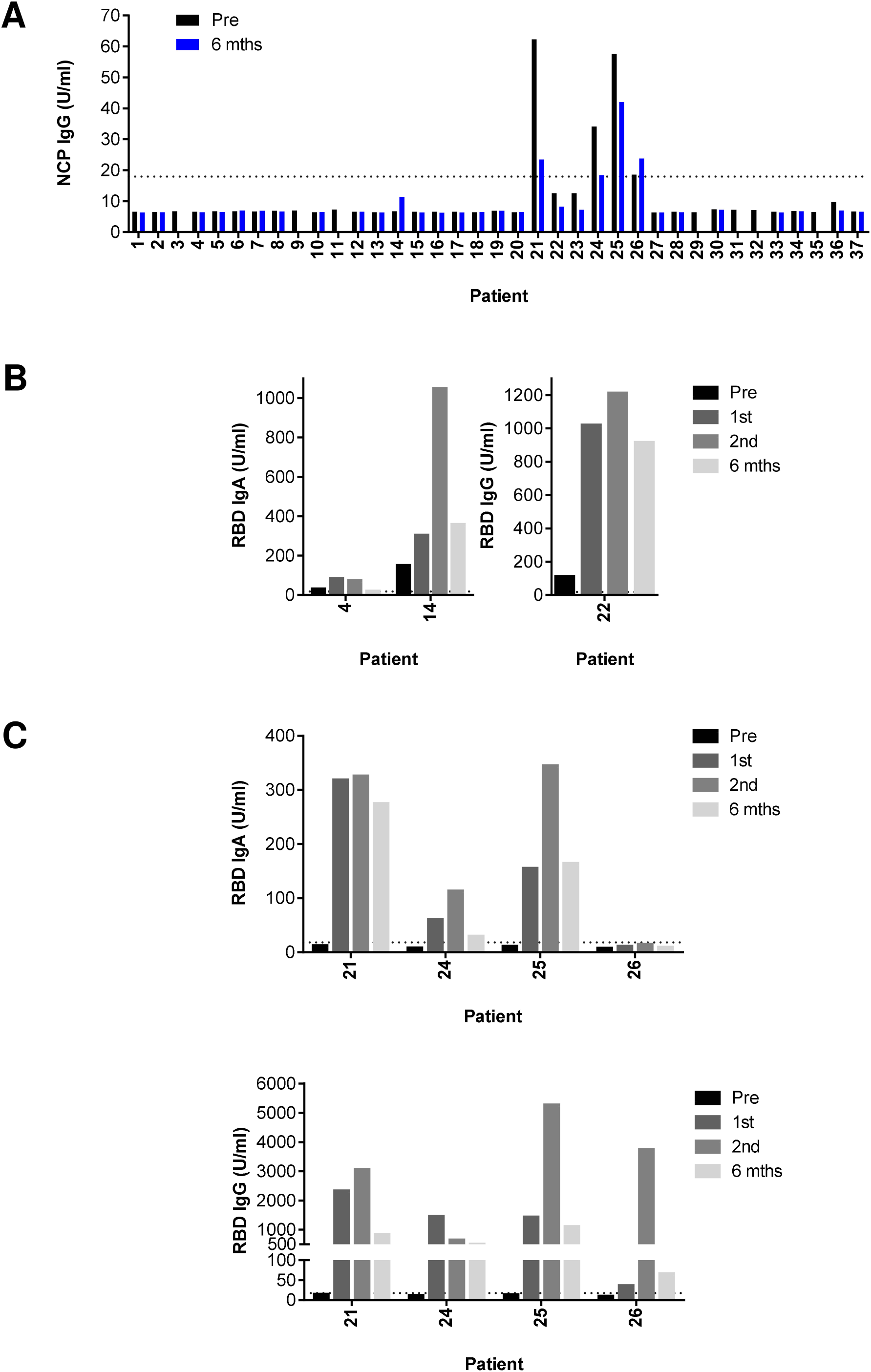
Serum levels of anti-NCP or anti-RBD antibodies. (**A**) Serum levels (U/ml) of anti-NCP IgG antibodies before the first vaccine dose (Pre), and 6 months (6 mths) after the second vaccine dose. (**B**) Serum levels (U/ml) of anti-RBD antibodies before the first (Pre) and second (1st) vaccine dose, and 4 weeks (2nd) and 6 months (6 mths) after the second vaccine dose in patients with pre-vaccination-elevated anti-RBD IgA (*left panel*) or IgG (*right panel*) antibodies. (**C**) Serum levels (U/ml) of anti-RBD IgA (*top panels*) or IgG (*bottom panels*) antibodies before the first (Pre) and second (1st) vaccine dose, and 4 weeks (2nd) and 6 months (6 mths) after the second vaccine dose in patients with post-vaccination-elevated anti-NCP IgG antibodies in **A**. In **A**–**C**, the dotted line indicates the value 18 U/ml. Values above 18 U/ml are positive.

**Figure S2.**
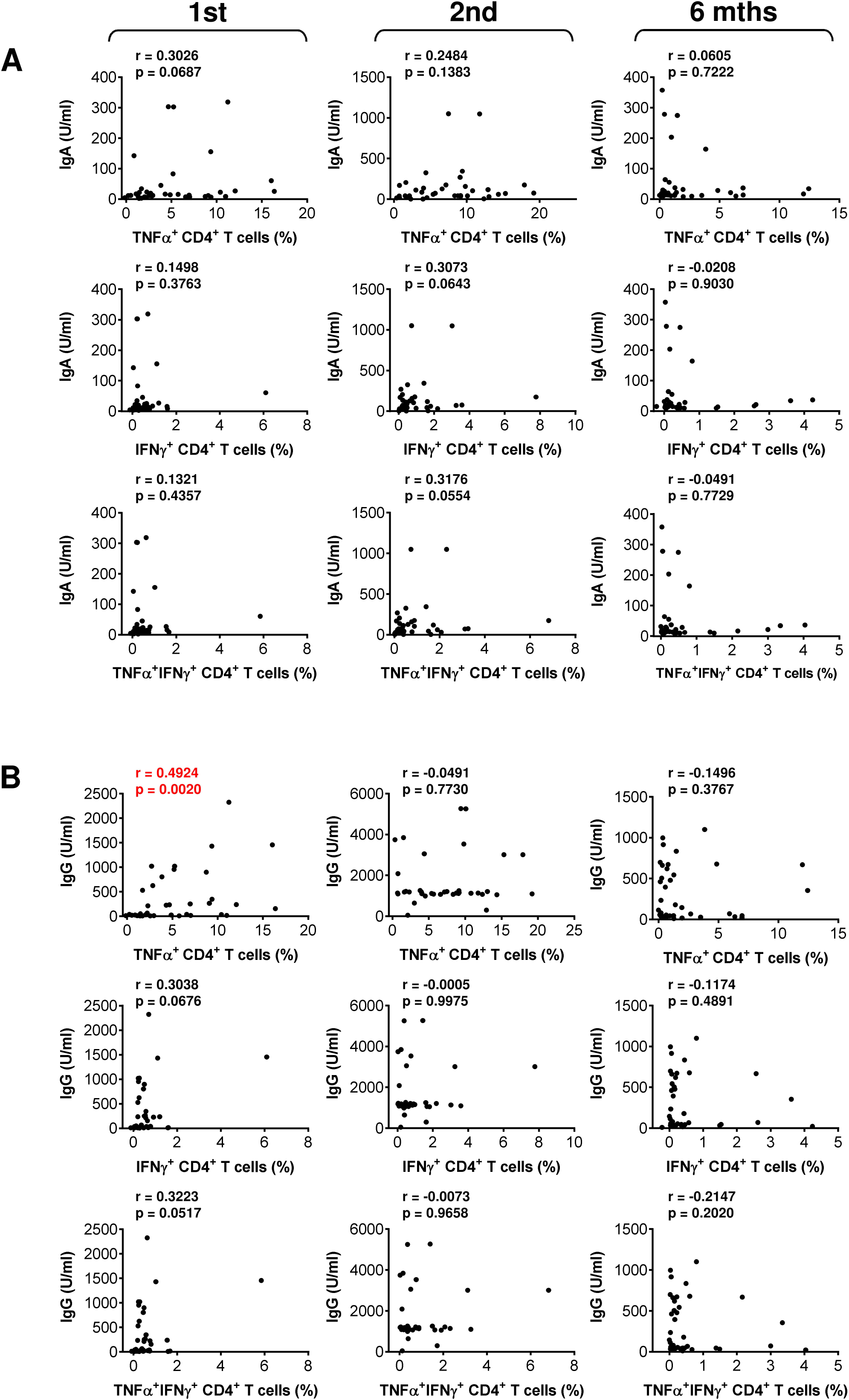
The association between SARS-CoV-2 spike glycoprotein peptide-reactive CD4^+^ T cells and serum levels of anti-RBD antibodies during and after the vaccination. (**A**–**B**) The correlations between the frequency of peptide-reactive TNFα-, IFNγ-, or TNFα/IFNγ-producing CD4^+^ T cells and serum levels (U/ml) of anti-RBD IgA (**A**) and IgG (**B**) antibodies before the second (1st) vaccine dose, and 4 weeks (2nd) and 6 months (6 mths) after the second vaccine dose. The Spearman’s rank-order correlation coefficient (r) and the significance (*p*-value; *n* = 37) are indicated.

**Figure S3.**
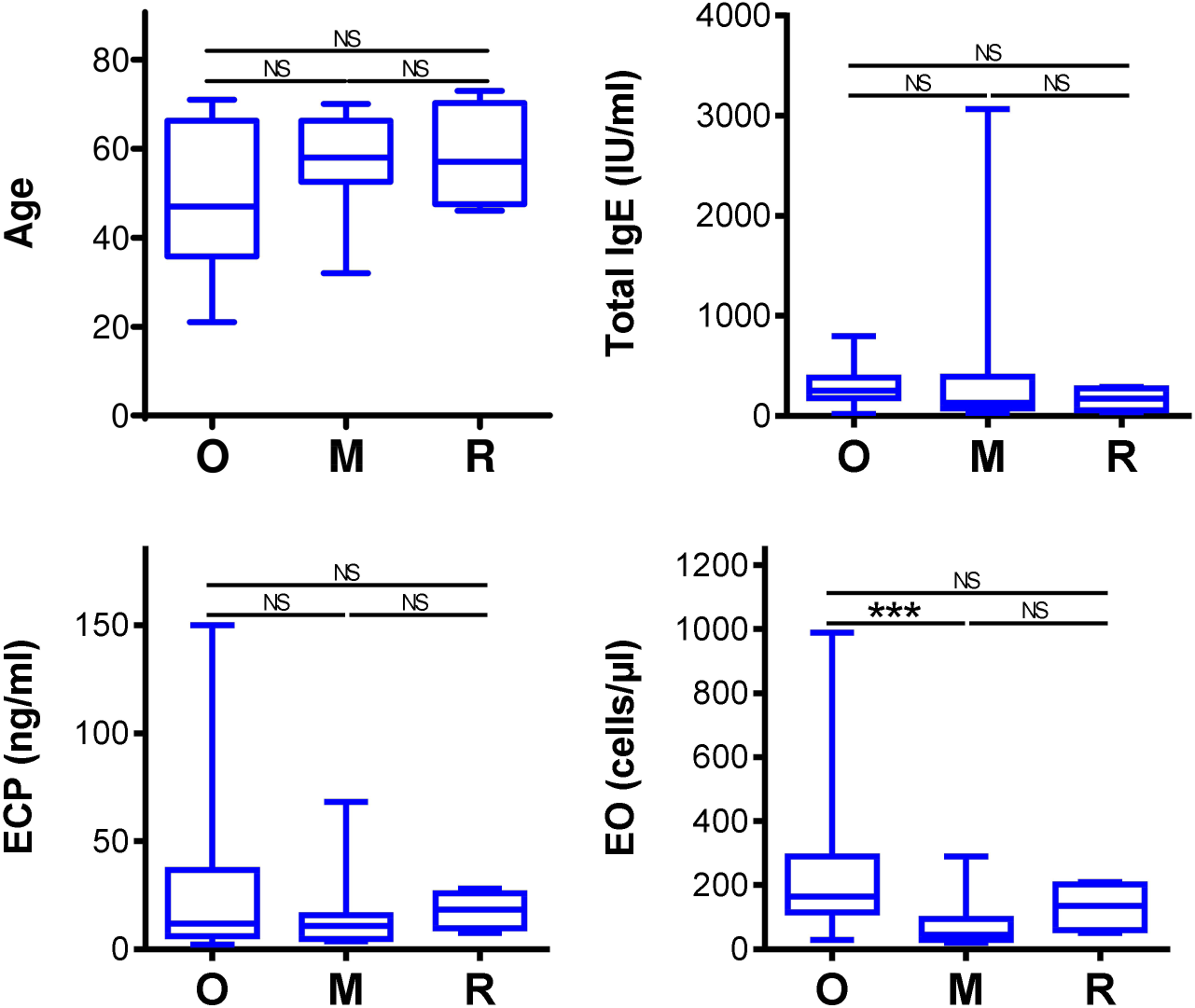
Demographics and clinical parameters of the biological therapy-stratified groups of patients. The patients were stratified into 3 groups based on the type of biological therapy. The groups were 18 patients on anti-IgE (O; omalizumab), 14 patients on anti-IL5 (M, mepolizumab), and 4 patients on anti-IL5 (R, reslizumab) therapy. The graphs with the age of patients (*top left*), total IgE serum levels (IU/ml) (*top right*), ECP (ng/ml) (*bottom left*), blood eosinophil counts (cells/µl) (*bottom right*) in the stratified groups are shown. Shown are box and whisker plots (2.5–97.5 percentile), and significances of differences among the groups (O, M, R) are indicated (^NS^*p>*0.05; *n* = 18 (O), 14 (M), or 4 (R); matched-pair one-way ANOVA with the Dunn’s posttest).

**Figure S4.**
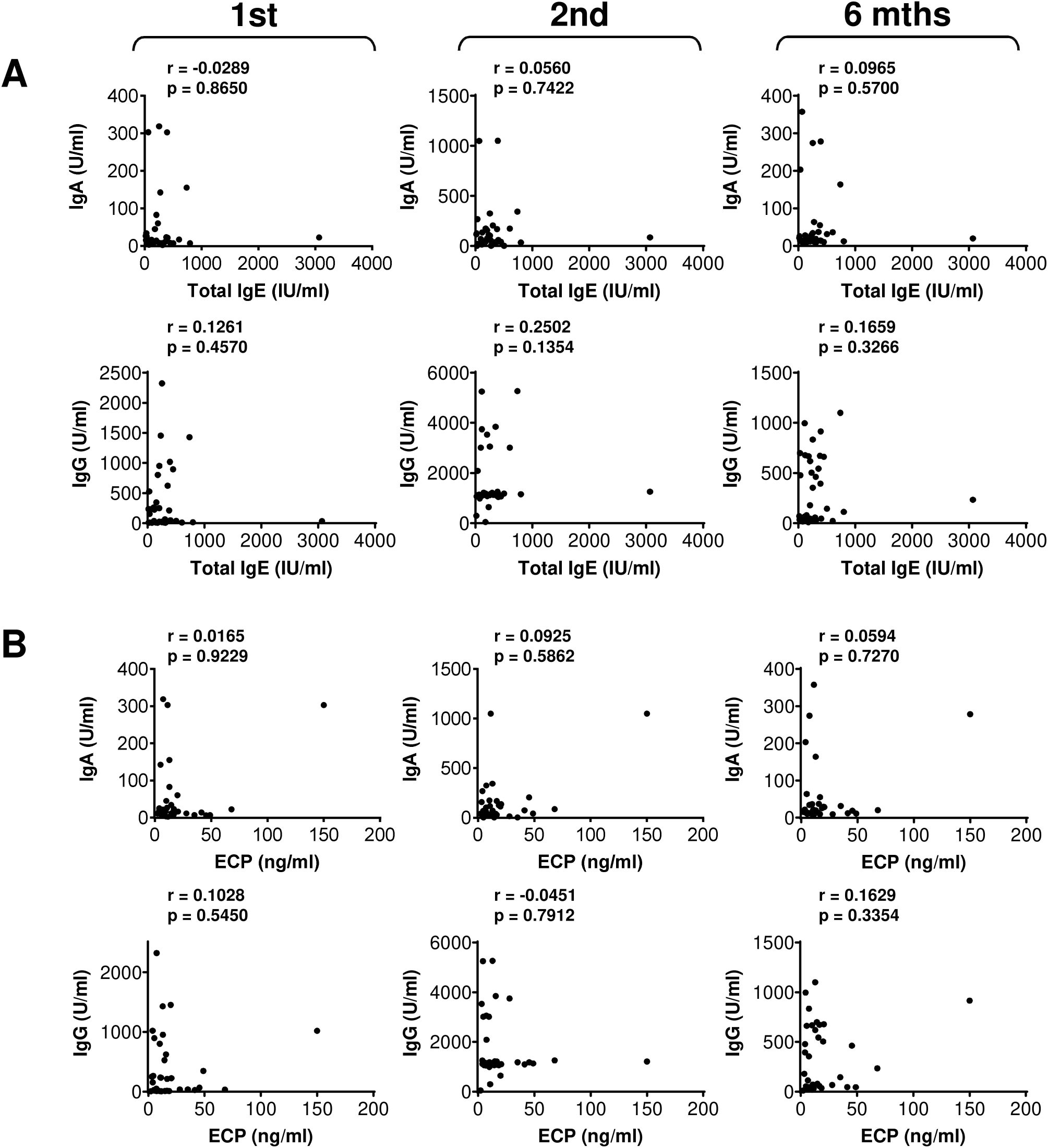
The association between clinical parameters (total IgE and ECP) and serum levels of anti-RBD antibodies during and after the vaccination. (**A**) The correlations between total IgE serum levels (IU/ml) and serum levels (U/ml) of anti-RBD IgA (*top panels*) and IgG (*bottom panels*) antibodies before the second (1st) vaccine dose, and 4 weeks (2nd) and 6 months (6 mths) after the second vaccine dose. (**B**) The correlations between ECP (ng/ml) and serum levels (U/ml) of anti-RBD IgA (*top panels*) and IgG (*bottom panels*) antibodies before the second (1st) vaccine dose, and 4 weeks (2nd) and 6 months (6 mths) after the second vaccine dose. In **A**–**B**, Spearman’s rank-order correlation coefficient (r) and the significance (*p*-value; *n* = 37) are indicated.

**Figure S5.**
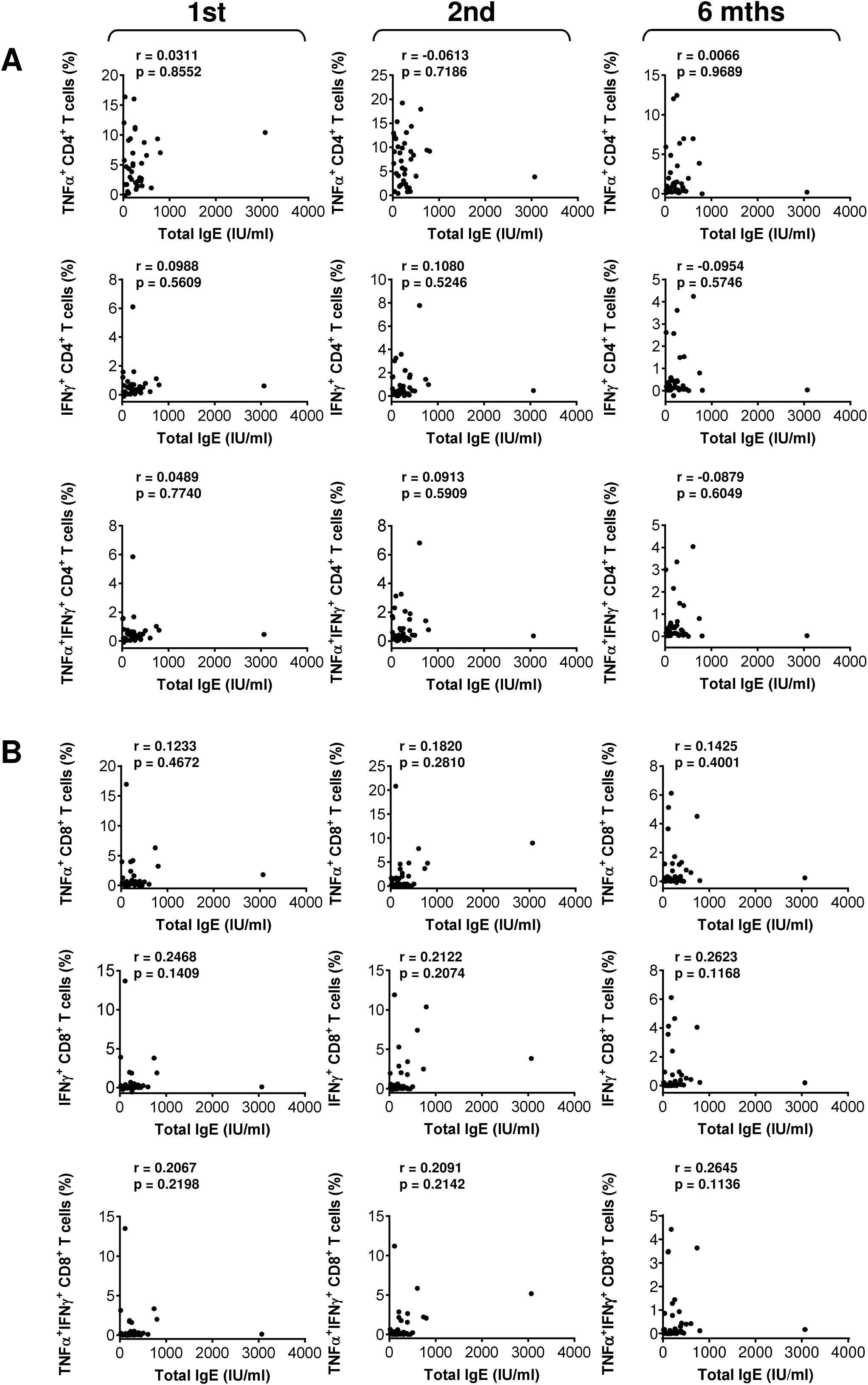
The association between total IgE serum levels and SARS-CoV-2 spike glycoprotein peptide-reactive T cells during and after the vaccination. (**A**–**B**) The correlations between total IgE serum levels (IU/ml) and TNFα-, IFNγ-, or TNFα/IFNγ-producing CD4^+^ (**A**) and CD8^+^ (**B**) T cells before the second (1st) vaccine dose, and 4 weeks (2nd) and 6 months (6 mths) after the second vaccine dose. In **A**–**B**, Spearman’s rank-order correlation coefficient (r) and the significance (*p*-value; *n* = 37) are indicated.

**Figure S6.**
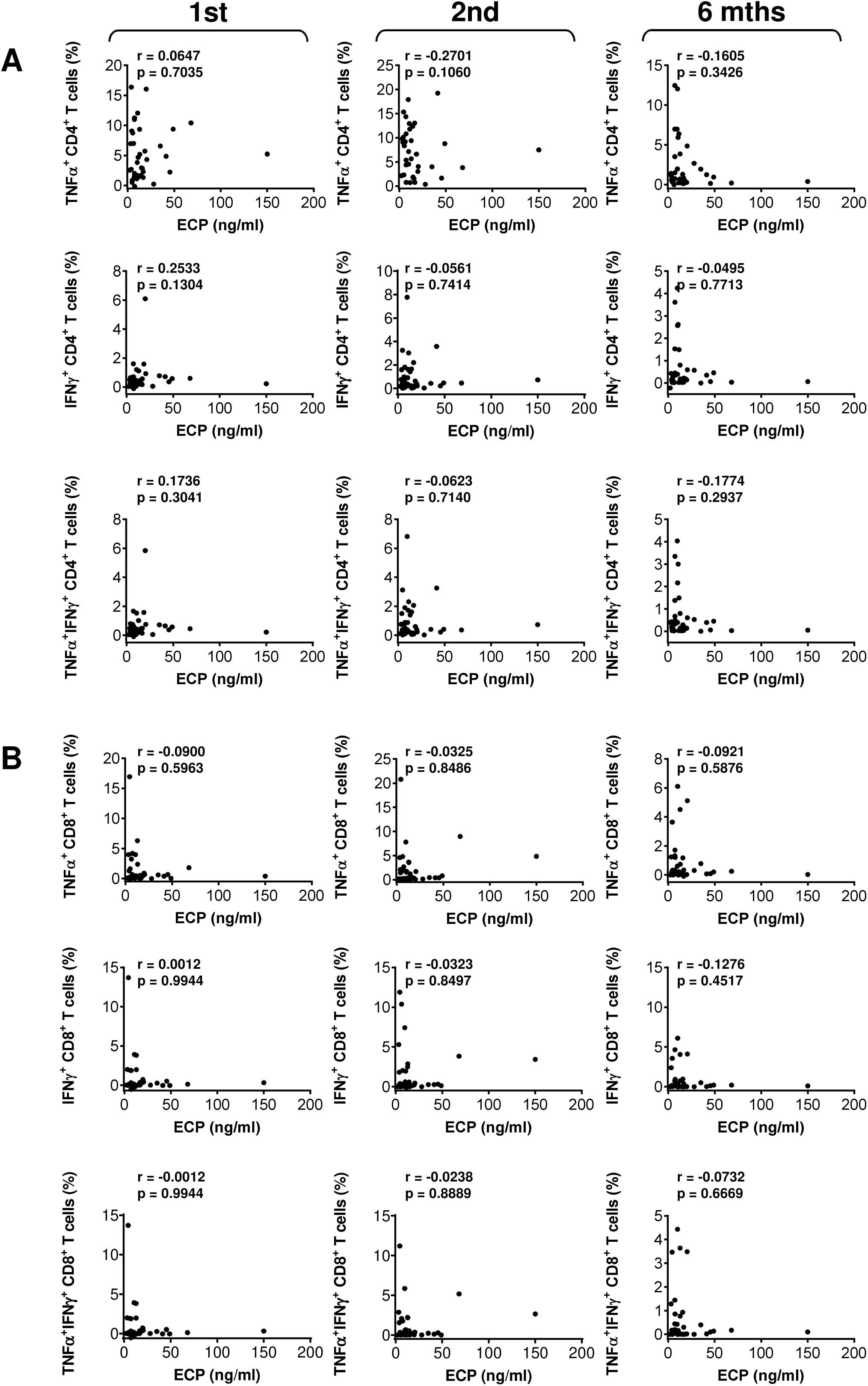
The association between ECP serum levels and SARS-CoV-2 spike glycoprotein peptide-reactive T cells during and after the vaccination. (**A**–**B**) The correlations between ECP (ng/ml) and TNFα-, IFNγ-, or TNFα/IFNγ-producing CD4^+^ (**A**) and CD8^+^ (**B**) T cells before the second (1st) vaccine dose, and 4 weeks (2nd) and 6 months (6 mths) after the second vaccine dose. In **A**–**B**, Spearman’s rank-order correlation coefficient (r) and the significance (*p*-value; *n* = 37) are indicated.

**Figure S7.**
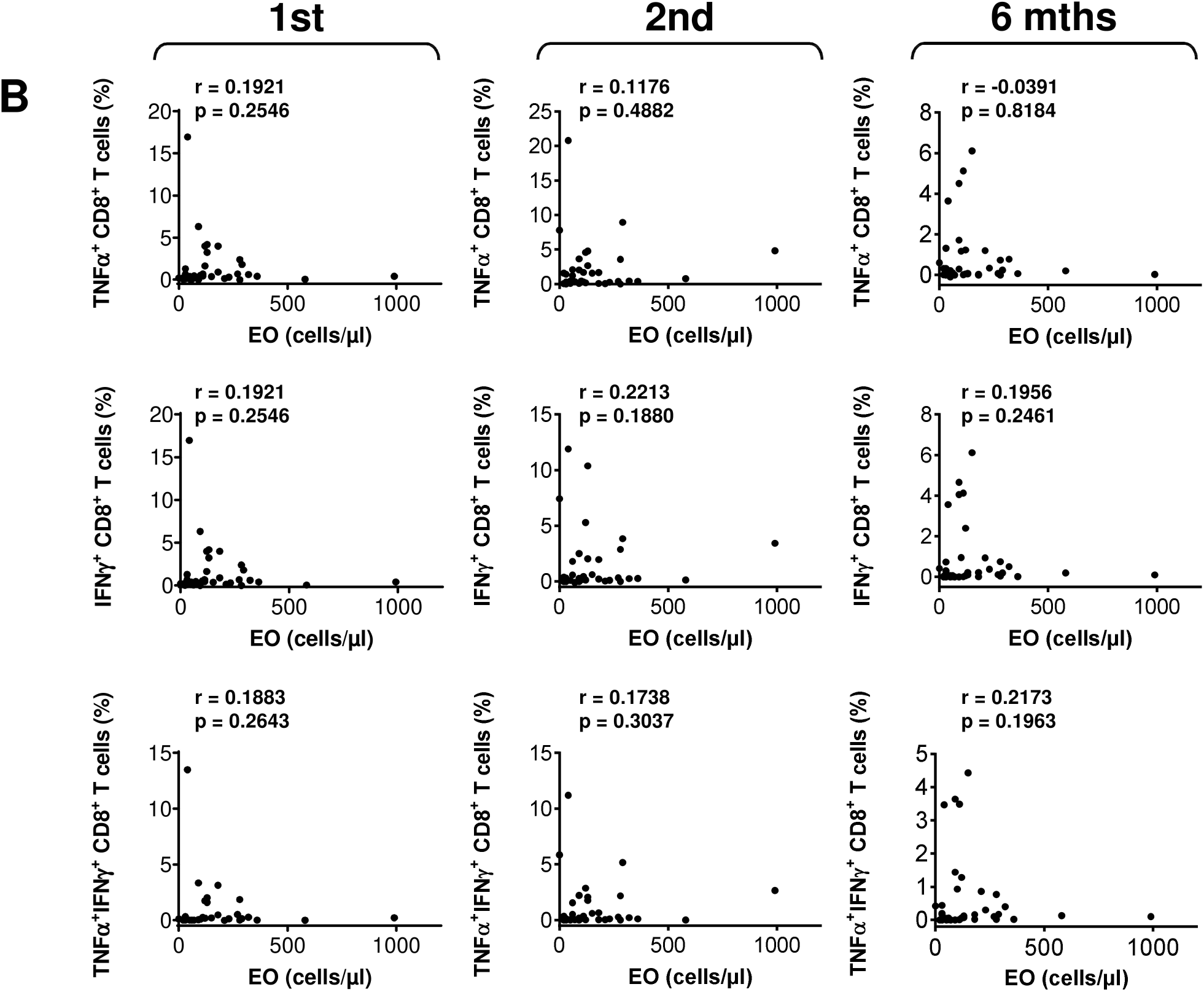
The association between blood eosinophil counts and SARS-CoV-2 spike glycoprotein peptide-reactive CD8^+^ T cells during and after the vaccination. (**A**–**B**) The correlations between blood eosinophil counts (cells/µl) and TNFα-, IFNγ-, or TNFα/IFNγ-producing CD8^+^ T cells before the second (1st) vaccine dose, and 4 weeks (2nd) and 6 months (6 mths) after the second vaccine dose. The Spearman’s rank-order correlation coefficient (r) and the significance (*p*-value; *n* = 37) are indicated.

**Figure S8.**
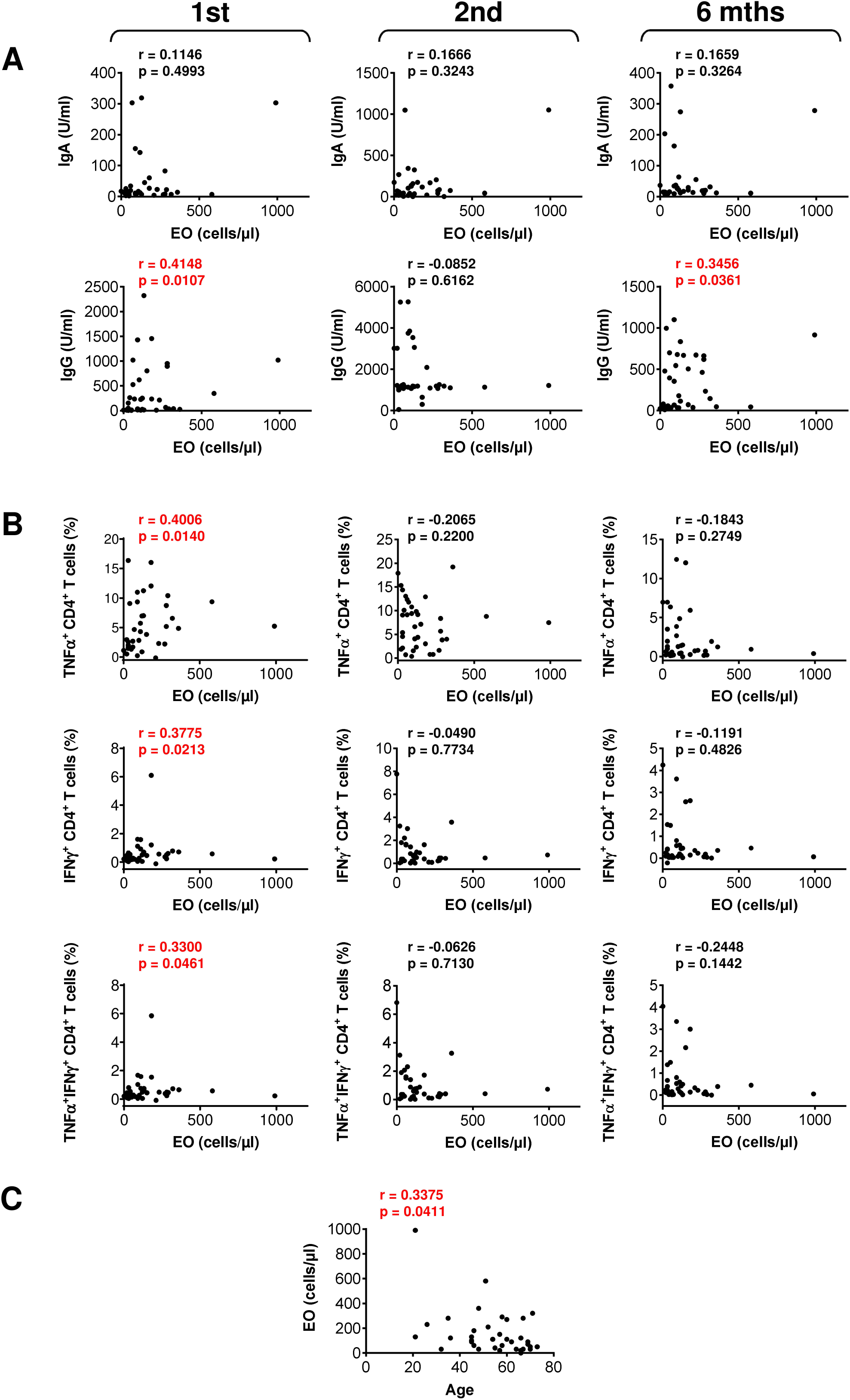
The impact of blood eosinophil counts on serum levels of anti-RBD antibodies, SARS-CoV-2 spike glycoprotein peptide-reactive CD4^+^ T cells and the patients’ age during and after the vaccination. (**A**–**B**) The correlations between blood eosinophil counts (cells/µl) and serum levels (U/ml) of anti-RBD IgA (*top panels*) and IgG (*bottom panels*) antibodies (**A**), or TNFα-, IFNγ-, or TNFα/IFNγ-producing CD4^+^ T cells (**B**) before the second (1st) vaccine dose, and 4 weeks (2nd) and 6 months (6 mths) after the second vaccine dose. (**C**) The correlation between blood eosinophil counts (cells/µl) and the patients’ age. In **A**–**C**, Spearman’s rank-order correlation coefficient (r) and the significance (*p*-value; *n* = 37) are indicated.

**Figure S9.**
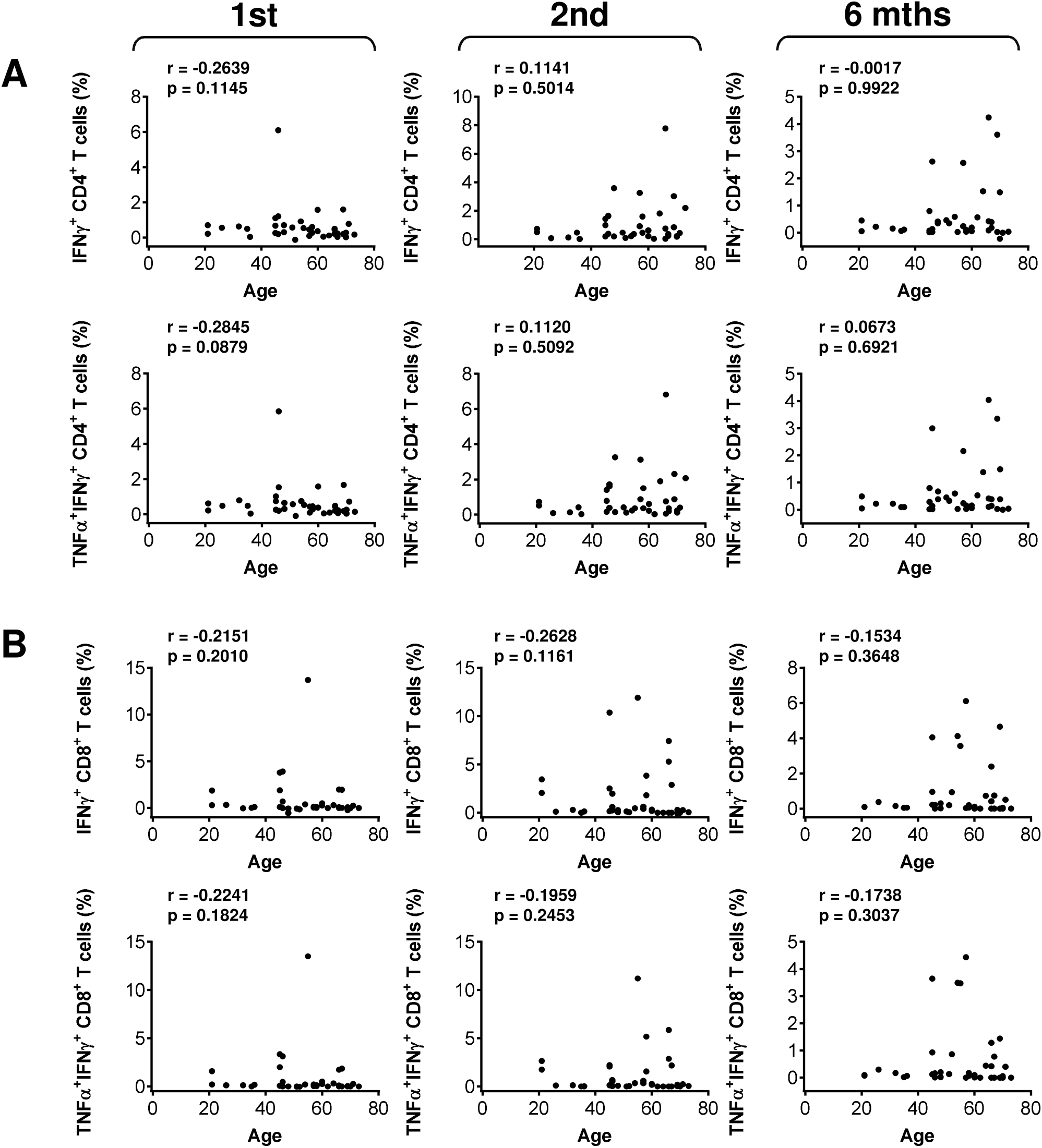
The impact of the patients’ age on SARS-CoV-2 spike glycoprotein peptide-reactive IFNγ- or TNFα/IFNγ-producing T cells during and after the vaccination. (**A**) The correlations between the patients’ age and the frequency of peptide-reactive IFNγ- (*top panels*) or TNFα/IFNγ- (*bottom panels*) producing CD4^+^ (**A**) and CD8^+^ (**B**) T cells before the second (1st) vaccine dose, and 4 weeks (2nd) and 6 months (6 mths) after the second vaccine dose. In **A**–**B**, Spearman’s rank-order correlation coefficient (r) and the significance (*p*-value; *n* = 37) are indicated.

## Notes

### Author Declarations

Ethics Committee/IRB of the Motol University Hospital in Prague gave ethical approval for this work. The protocol approval number is EK-346/21.

